# Age-dependent assessment of genes involved in cellular senescence, telomere and mitochondrial pathways in human lung tissue of smokers, COPD and IPF: Associations with SARS-CoV-2 COVID-19 ACE2-TMPRSS2-Furin-DPP4 axis

**DOI:** 10.1101/2020.06.14.20129957

**Authors:** Krishna P. Maremanda, Isaac K. Sundar, Dongmei Li, Irfan Rahman

**Affiliations:** Department of Environmental Medicine, University of Rochester Medical Center, Rochester, NY, USA; Department of Clinical & Translational Research, University of Rochester Medical Center, Rochester, NY, USA

## Abstract

Aging is one of the key contributing factors for chronic obstructive pulmonary diseases (COPD) and other chronic inflammatory lung diseases. Cigarette smoke is a major etiological risk factor that has been shown to alter cellular processes involving mitochondrial function, cellular senescence and telomeric length. Here we determined how aging contribute to the alteration in the gene expression of above mentioned cellular processes that play an important role in the progression of COPD and IPF. We hypothesized that aging may differentially alter the expression of mitochondrial, cellular senescence and telomere genes in smokers and patients with COPD and IPF compared to non-smokers. Total RNA from human lung tissues from non-smokers, smokers, and patients with COPD and IPF were processed and analyzed based on their ages (younger: <55 yrs and older: >55 yrs). NanoString nCounter panel was used to analyze the gene expression profiles using a custom designed codeset containing 112 genes including 6 housekeeping controls (mitochondrial biogenesis and function, cellular senescence, telomere replication and maintenance). mRNA counts were normalized, log2 transformed for differential expression analysis using linear models in the *limma* package (R/Bioconductor). Data from non-smokers, smokers and patients with COPD and IPF were analyzed based on the age groups (pairwise comparisons between younger vs. older groups). Several genes were differentially expressed in younger and older smokers, and patients with COPD and IPF compared to non-smokers which were part of the mitochondrial biogenesis/function (*HSPD1, FEN1, COX18, COX10, UCP2 & 3*), cellular senescence (*PCNA, PTEN, KLOTHO, CDKN1C, TNKS2, NFATC1 & 2, GADD45A*) and telomere replication/maintenance (*PARP1, SIRT6, NBN, TERT, RAD17, SLX4, HAT1*) target genes. Interestingly, *NOX4* and *TNKS2* were increased in the young IPF as compared to the young COPD patients. Genes in the mitochondrial dynamics and other quality control mechanisms like *FIS1* and *RHOT2* were decreased in young IPF compared to their age matched COPD subjects. ERCC1 (Excision Repair Cross-Complementation Group 1) and *GADD45B* were higher in young COPD as compared to IPF. Aging plays an important role in various infectious diseases. Elderly patients with chronic lung disease and smokers were found to have high incidence and mortality rates in the current pandemic of SARS-CoV-2 infection. Immunoblot analysis in the lung homogenates of smokers, COPD and IPF subjects revealed increased protein abundance of important proteases and spike proteins like TMPRSS2, furin and DPP4 in association with a slight increase in SARS-CoV-2 receptor ACE2 levels. This may further strengthen the observation that smokers, COPD and IPF subjects are more prone to COVID-19 infection. Overall, these findings suggest that altered transcription of target genes that regulate mitochondrial function, cellular senescence, and telomere attrition add to the pathobiology of lung aging in COPD and IPF and other smoking-related chronic lung disease in associated with alterations in SARS-CoV-2 ACE2-TMPRSS2-Furin-DPP4 axis for COVID-19 infection.

## Introduction

Aging is an important factor influencing the overall lung health and function ^1,2^. Lung function declines with progress in age after lung maturation. Evidences suggest that a significant contribution of various environmental factors influence the ageing lung ^3^. According to the Behavioral Risk Factor Surveillance System (BRFSS) data from 2017, 6.2% (age-adjusted) US adults were reported to have chronic obstructive pulmonary disease (COPD) ^4^. Further, there has been an increasing reports of young COPD population visiting for different hospital services ^5^. Ageing influences many chronic lung diseases like COPD, idiopathic pulmonary fibrosis (IPF) and asthma. Also chronic lung diseases like asthma and IPF share some of the common yet distinct features compared to COPD ^6^. Environmental stress factors like smoking remains a common influencing factor for the disease progression in all these three cases.

Cigarette smoke (CS) is one of the strongest contributing risk factors in the pathogenesis of COPD along with the decline in lung function ^7^. Cellular senescence is a process of irreversible cell cycle arrest, having both beneficial and harmful effects depending on the cell state ^8^. CS plays a role in advancing the lung aging by altering the process of cellular senescence ^9^. Several factors including oxidative stress influence the process of cellular senescence. Telomeres and mitochondria play a major role in influencing the process of cellular senescence and are often associated with maintaining the lung health ^8,10,11^. Earlier reports from our laboratory and others have shown that smoking and COPD is associated with the mitochondrial damage and dysfunction, altering the process of cellular metabolism and function ^12,13^. Similarly, telomere dysfunction was also associated with smoking and COPD ^14,15^. Mitochondrial and telomere dysfunction also play a causative role in the progression of IPF ^16-18^.

Several molecular mechanisms were identified and reported in relation to CS causing COPD and associated complications ^19^. Cigarette smoke alters several key functions in the cells, among them the crucial genes related to mitochondrial function, cellular senescence and telomeric length were selected in the current study to observe for any differential changes among young and old age groups categorized as non-smokers, smokers and COPD groups. Our previous studies showed independent contributions of these canonical signaling pathways and how they contribute towards the development of premature lung aging in chronic lung diseases, such as COPD/emphysema ^20-24^. Accumulating evidence suggest the close relationship of all these three pathways in influencing lung ageing and disease ^11,25,26^. Senescent cells are found in many age-related/chronic diseases ^27^. Studies from our laboratory showed that mice from different age group when exposed to chronic air and CS influence the process of lung inflammation and senescence. Chronic CS exposure in lung epithelial cells and mice increases several markers of cellular senescence ^9,28^. Recently, it was reported that serum from COPD patients can induce senescence in lung epithelial cells ^29^, giving strength to the importance of this area that needs to be explored further. Several DNA damaging agents occur in smoke, which may activate of DNA damage response by influencing telomere function over the time and tend to accumulate senescent cells. However, recent meta-analysis suggests that even though smokers are associated with shorter telomere length, the study implicates that smoking does not accelerate the telomere attrition in leucocytes ^30^.

Smoking and COPD conditions altered the expressions of many key genes involved in the above three crucial pathways of cellular maintenance. The current study was undertaken to determine the changes in genes related to mitochondrial biogenesis and function, telomere function and cellular senescence with respect to their age in human lungs. This study is important in unraveling some of the potential biomarkers to differentiate and follow the course of ageing in COPD. Keeping in view the importance of these genes in both COPD and IPF, we have also made comparisons between the similar age grouped COPD and IPF subjects, using the same set of gene panels.

Aging is one of the key components, which decides the subject’s susceptibility to various diseases and infections ^31^. Inflammaging/immunosenescence defines the condition to combat various illness. Recent pandemic of Severe Acute Respiratory Syndrome (ARDS) Coronavirus 2 (SARS-CoV-2) was thought to affect more elderly people especially men ^32-34^. Men with age-related comorbidities has higher mortality rate during Coronavirus Disease 2019 (COVID-19)^35^. Comorbidities like cardiovascular disease, diabetes and chronic respiratory diseases present a high mortality rate^36^. The studies involving the role of lung aging and senescence play an important role in understanding the role of multiple players involved in combating against SARS-CoV-2 infection and can lead to potential druggable targets to combat it. Considering the important role played by some of the crucial receptors and targets in COVID-19 ^37^, we determined the protein expression of SARS-CoV-2 receptor Angiotensin Converting Enzyme 2 (ACE2) and aiding proteases like transmembrane protease serine protease-2 (TMPRSS-2) and spike protein furin in the lung homogenates of non-smokers, smokers, COPD and IPF subjects. We have also determined for the levels of dipeptidyl peptidase 4 (DPP4), which acts as a receptor for the similar class of coronavirus which is Middle East Respiratory Syndrome Coronavirus, (MERS-CoV)^38^.

## Materials and Methods

### Scientific rigor and reproducibility

Rigorous and unbiased approach were used to ensure full and detailed reporting of both methods and analyzed data.

### Ethical approval: Institutional biosafety an review board approvals

#### Ethics statement

The current study was approved for the procurement of the human lung tissues as de-identified tissues by the Materials Transfer Agreement and Procurement (Institutional Review Board), and laboratory protocols by the Institutional Biosafety Committee of the University of Rochester Medical Center, Rochester, NY. Patients’ data or patients are not directly involved in this study as the lung tissues were procured from several agencies (see below). All patients/subjects were of age 21 and above. All methods were carried out in accordance with relevant guidelines and regulations of the University of Rochester, Rochester, NY.

### Human lung tissues

The human peripheral lung tissues from non-smokers, smokers and COPD were procured/obtained from the NDRI (National Disease Research Interchange; the samples were collected from patients with various cause of deaths reported such as cardiac arrest or trauma/accidents, for most of the samples lower peripheral lung lobes were used or as supplied), LTRC (Lung Tissue Research Consortium of the NHLBI) and Department of Medicine and Pathology, and of the University of Helsinki Hospital, Finland as described in our previous reports ^39^. The clinical characteristics of the subjects used in the current study are given in **Table 1**. The subjects were broadly classified into two age groups: young age (≤55 years) group, old age (>55 years) group, as per previous studies ^40^. Although there were some co-morbid conditions reported for the specimens from COPD patients (which were on various medications), the tissues were assigned to different groups based on the age, smoking status and lung disease status (normal vs smoker’s normal vs COPD) reported during the procurements of the specimens. Even though the definitions across various reports vary, the samples in the non-smokers group were either reported to have no smoking history or doesn’t fit under current smokers with a long past tobacco use specially in old non-smokers, but have normal lungs, smokers have current smoking history with lung function, and COPD have diseased lungs classified as COPD. Additional comparisons were made between COPD (8 additional samples from above groups were added in addition to the 24 samples mentioned in **Table 1**) and IPF lung samples (3 in young IPF, 13 in old IPF) based on their age to check for the changes in the same custom gene panel (**Tables 2 and 3**).

**Table 1.**
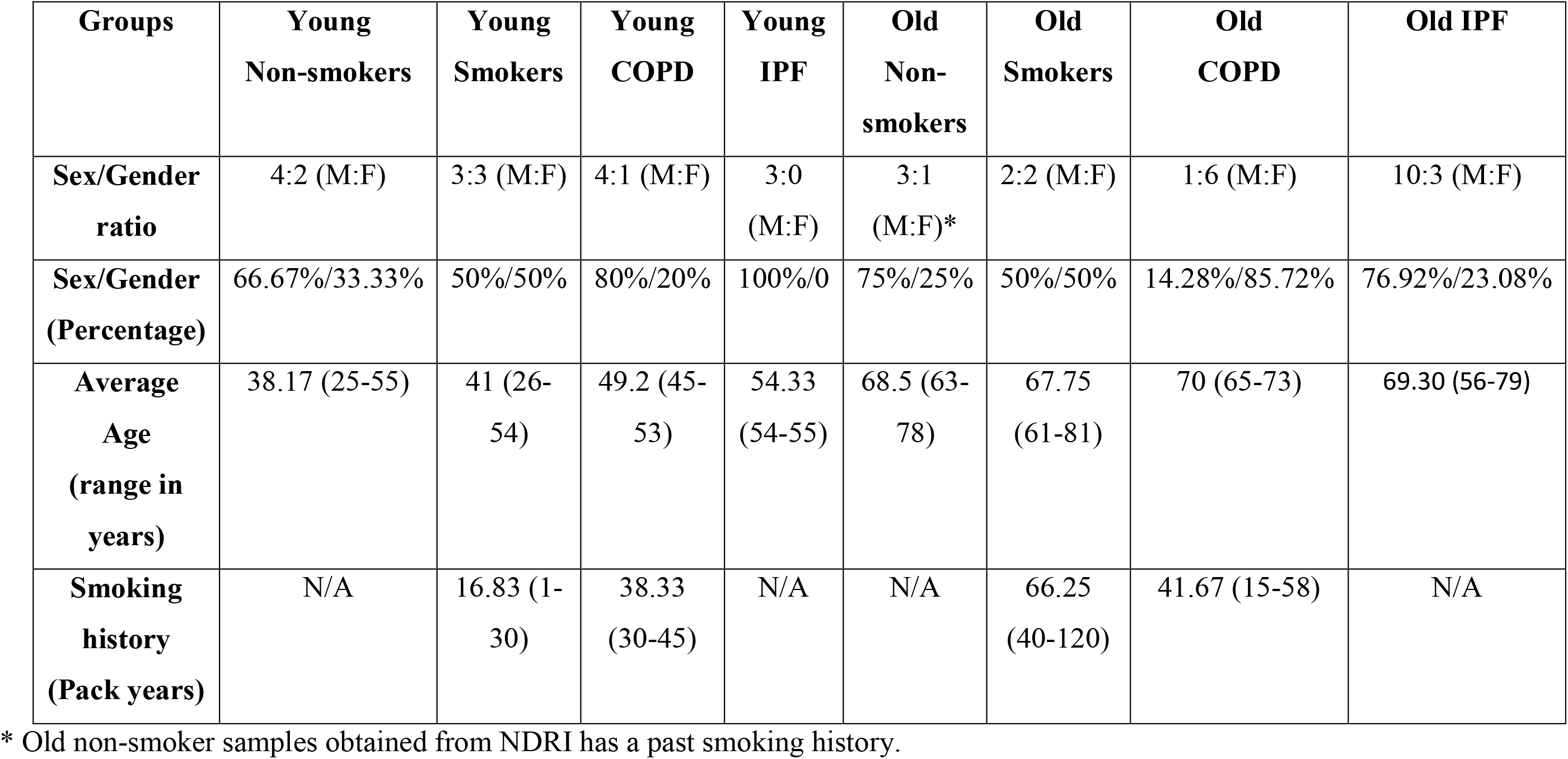
Clinical characteristics of non-smokers, smokers, and COPD and IPF subjects/patients

| **Groups** | **Young**<br>**Non-smokers** | **Young**<br>**Smokers** | **Young**<br>**COPD** | **Young**<br>**IPF** | **Old**<br>**Non-**<br>**smokers** | **Old**<br>**Smokers** | **Old**<br>**COPD** | **Old IPF** |
| --- | --- | --- | --- | --- | --- | --- | --- | --- |
| **Sex/Gender**<br>**ratio** | 4:2 (M:F) | 3:3 (M:F) | 4:1 (M:F) | 3:0<br>(M:F) | 3:1<br>(M:F)* | 2:2 (M:F) | 1:6 (M:F) | 10:3 (M:F) |
| **Sex/Gender**<br>**(Percentage)** | 66.67%/33.33% | 50%/50% | 80%/20% | 100%/0 | 75%/25% | 50%/50% | 14.28%/85.72% | 76.92%/23.08% |
| **Average**<br>**Age**<br>**(range in**<br>**years)** | 38.17 (25-55) | 41 (26-54) | 49.2 (45-53) | 54.33<br>(54-55) | 68.5 (63-78) | 67.75<br>(61-81) | 70 (65-73) | 69.30 (56-79) |
| **Smoking**<br>**history**<br>**(Pack years)** | N/A | 16.83 (1-30) | 38.33<br>(30-45) | N/A | N/A | 66.25<br>(40-120) | 41.67 (15-58) | N/A |

\* Old non-smoker samples obtained from NDRI has a past smoking history.

**Table 2.** Altered genes in comparisons between Young Nonsmokers and Young IPF subjects

| List of genes that were found to be increased in young IPF compared to young Nonsmokers |  |
| --- | --- |
| Genes | p values |
| AIFM2 | 0.027262 |
| COX10 | 0.008096 |
| IGF1 | 0.017368 |
| RAD50 | 0.041025 |
| List of genes that were found to be decreased in young IPF compared to young Nonsmokers |  |
| AIP | 0.007814 |
| AKT1 | 0.028541 |
| ATP5O | 0.010191 |
| CDK2 | 0.001304 |
| CDKN1B | 0.034287 |
| ERCC1 | 0.043178 |
| FIS1 | 0.016032 |
| GADD45B | 0.000329 |
| GSK3B | 0.000953 |
| HNRNPD | 0.035938 |
| ID1 | 0.007673 |
| IGF1R | 0.017547 |
| NFATC1 | 0.028756 |
| NFATC2 | 0.015133 |
| RAP1A | 0.014367 |
| RAPGEF1 | 0.025203 |
| RELA | 0.014566 |
| SP1 | 0.024547 |
| TGFB1 | 0.017524 |
| TIMM10B | 0.011584 |
| UXT | 0.034998 |

**Table 3.** Altered genes in comparisons between Old Nonsmokers and Old IPF subjects. List of genes that were found to be increased in old IPF compared to old Nonsmokers

| Genes | p values |
| --- | --- |
| AIFM2 | 0.036937 |
| ALDH1A3 | 0.004268 |
| BAK1 | 0.002375 |
| FEN1 | 0.017398 |
| PARP1 | 0.029557 |
| PCNA | 0.001159 |
| RPA2 | 0.022724 |
| TERF2 | 0.013038 |
| List of genes that were found to be decreased in old IPF compared to old Nonsmokers |  |
| CDK2 | 0.016647 |
| ERCC1 | 0.007071 |
| FOXO1 | 0.04744 |
| ID1 | 0.03526 |
| IGF1R | 0.014197 |

### RNA isolation from lung tissues

Total RNA was extracted from the human lung tissues stored at -80°C or in the RNA later, using Direct-Zol RNA miniprep plus kit (Zymo research, R2071) according to the manufacturer’s instructions. RNA concentration was measured using a Nanodrop 1000 (Thermo fisher scientific, USA). Various genes involved in Mitochondrial Biogenesis and Function, Telomere Replication and Maintenance and Cellular Senescence pathways were included in the custom code sets (Supplementary **Table S1)**. The code set contained a total of 112 genes including 6 reference genes (*Abcf1, Hprt, Polr1b, Rplp0, Ldha, Gusb*) for gene normalization (Supplementary **Table S2)**. The samples were processed through the NanoString nCounter system (NanoString Technologies Seattle, WA, USA). A total of 400ng RNA was submitted after adjusting the samples to a minimum of 20µg/µl as per the requirements.

### Validation of gene targets using quantitative real-time PCR

Selected mRNA, which were found to be significantly and differentially altered which were further validated for their expression using qPCR in samples (three samples per group) used in the study as described earlier ^41^. The primers were obtained from Bio-Rad, except for *FEN1* (F-CACCTGATGGGCATGTTCTAC, R-CTCGCCTGACTTGAGCTGT) and *18S* (F-GTAACCCGTTGAACCCCATT, R-CCATCCAATCGGTAGTAGCG), used as internal control were obtained from integrated DNA Technologies. Results were represented as pairwise comparisons to compliment the observations made by NanoString analysis. Student t-test was used to determine the level of significance between two groups, while ANOVA was used for multiple group comparisons.

### Western blot analysis in human lung homogenates

Total protein isolated from the lung homogenates of non-smokers, smokers, COPD and IPF were reduced and separated using the pre-made polyacrylamide gels (Bio-Rad) ^41^. The transferred membranes were probed for some of the important protein involved in the COVID-19, like tmprss2 (ab92323), furin (ab183495), DPP4 (ab28340) and ACE2 (ab108252). All the antibodies used in the current study was procured from the Abcam. The relative expression and equal loading as assessed using the Ponceau S staining or β-actin after stripping of the blots.

### Data processing and statistical analysis

Nanostring mRNA counts were first normalized using the *NanoStringNorm* function in the statistical analysis software R on log2 transformed data. Geometric mean of reference genes was used to remove the technique variation and background expression. Differential analysis was conducted using linear models in the *limma* package (R/Bioconductor) after adjusting for the gender difference. Comparison among different experimental groups were performed using linear contrasts within the linear model framework; moderated *t* statistics was used to determine the differences in the gene expression levels between groups with empirical Bayes approach. The Benjamini-Hochberg procedure was used to adjust the *p* values to control the false discovery rates at 5%. The analyzed data was represented in the graphs as y-axis showing the negative log10 P-value and x-axis representing log2 fold change across each pairwise comparisons as described previously ^42^. The significantly altered gene data were shown as dot plot representation in supplementary **Figures S1-9**. Four samples from each age group were used for comparisons among non-smokers, smokers and COPD groups (as given in Supplementary **Table S2**). Comparisons with IPF includes all the samples as mentioned in the **Table 1**.

## Results

Overall the study consisted of 24 lung tissue samples from different sources as mentioned above. The collected tissues were classified into six different groups based on age, the smoking and disease status. Further, comparative gene analysis was also done based on the smoking and disease status irrespective of the age. There were no genes in common that were changed in any of the comparisons involving all the three groups *i*.*e*. non-smokers, smokers and COPD. However, the individual group wise comparisons were reported here.

### Differentially expressed genes in young non-smokers versus young smokers versus young COPD groups

First, we analyzed differentially expressed transcript levels among young non-smokers vs. young smokers, young smokers vs. young COPD and young non-smokers vs. young COPD **(Figure 1)**. We found 5 genes were differentially expressed in young non-smokers vs. young smokers’ pairwise comparison. Out of 5 genes, the transcript levels of 4 genes (*NFATC1, NFATC2, GADD45A, and CDKN1A*) were decreased and 1 gene (*PARP1*) was increased in the young smokers as compared to young non-smokers group (**Figures 2 and 3 A**). Next, we compared genes differentially expressed in young smokers vs. young COPD pairwise comparison. Out of 5 genes, transcript levels of 1 gene (*SIRT6*) was decreased and the remaining 4 genes (*RAD17, CDKN1C, COX10* and *KLOTHO*) were significantly increased in young smokers as compared to young COPD group (**Figures 2A and 3B**). Finally, we found 6 genes differentially expressed among young non-smokers vs. young COPD pairwise comparison. Out of 6 genes, the transcript levels of 2 genes *CDKN1C* and *KLOTHO* that belong to cellular senescence panel were decreased in the young COPD as compared to young non-smokers group. While the transcript levels of remaining 4 genes *PARP1, SIRT6, TERT* and *SLX4* were increased in young COPD as compared to young non-smokers group (**Figures 2A and 3C**). Overall, 4 genes *PARP1, SIRT6, KLOTHO* and *CDKN1C* were among the common target genes that were differentially expressed in young COPD as compared to young non-smokers and young smokers groups.

**Figure 1.**
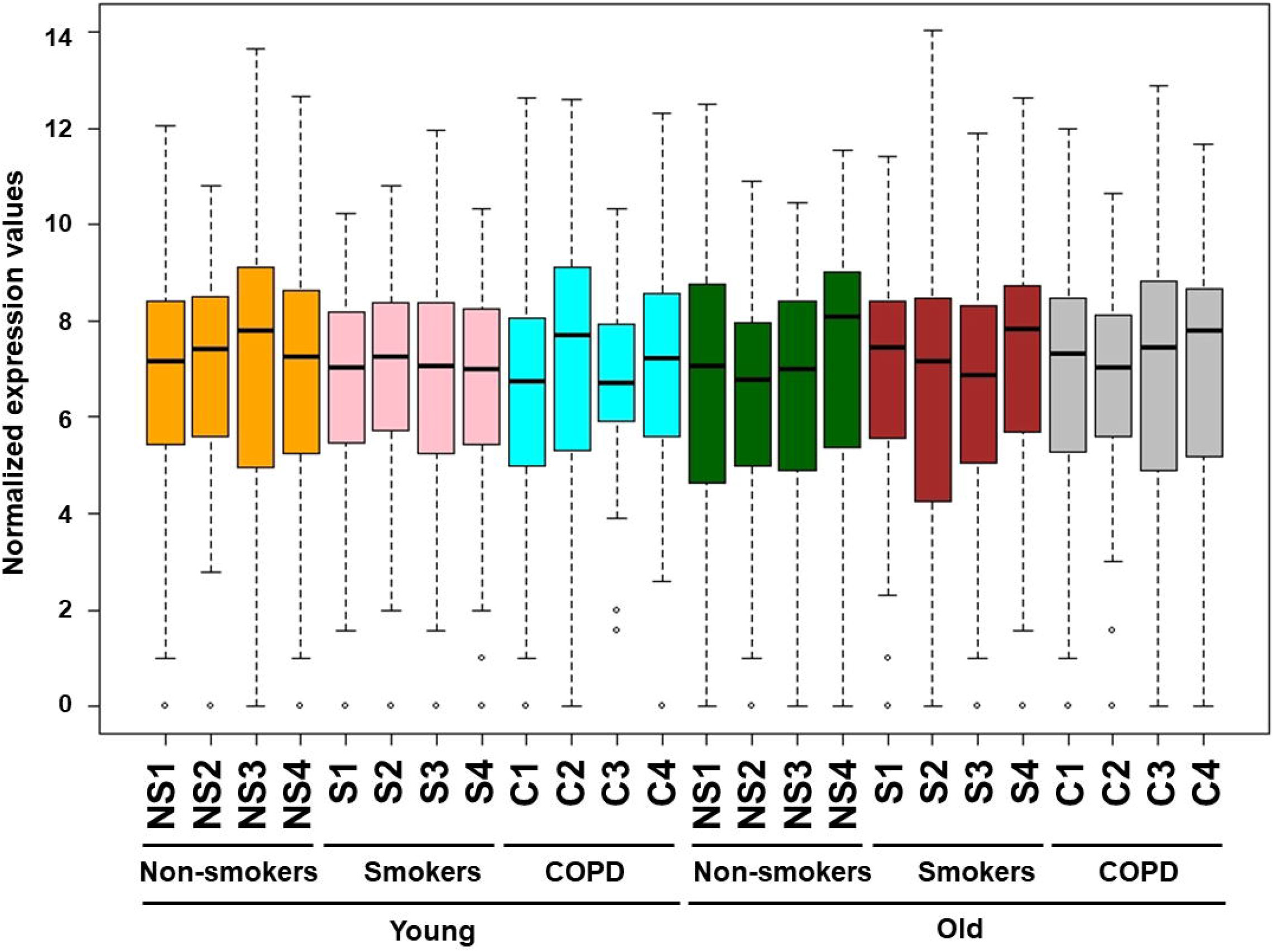
Boxplot analysis of normalized mRNA transcript analyzed by NanoString. Boxplot shows distribution of normalized gene expression levels from young and old non-smokers, smokers and COPD subjects.

**Figure 2.**
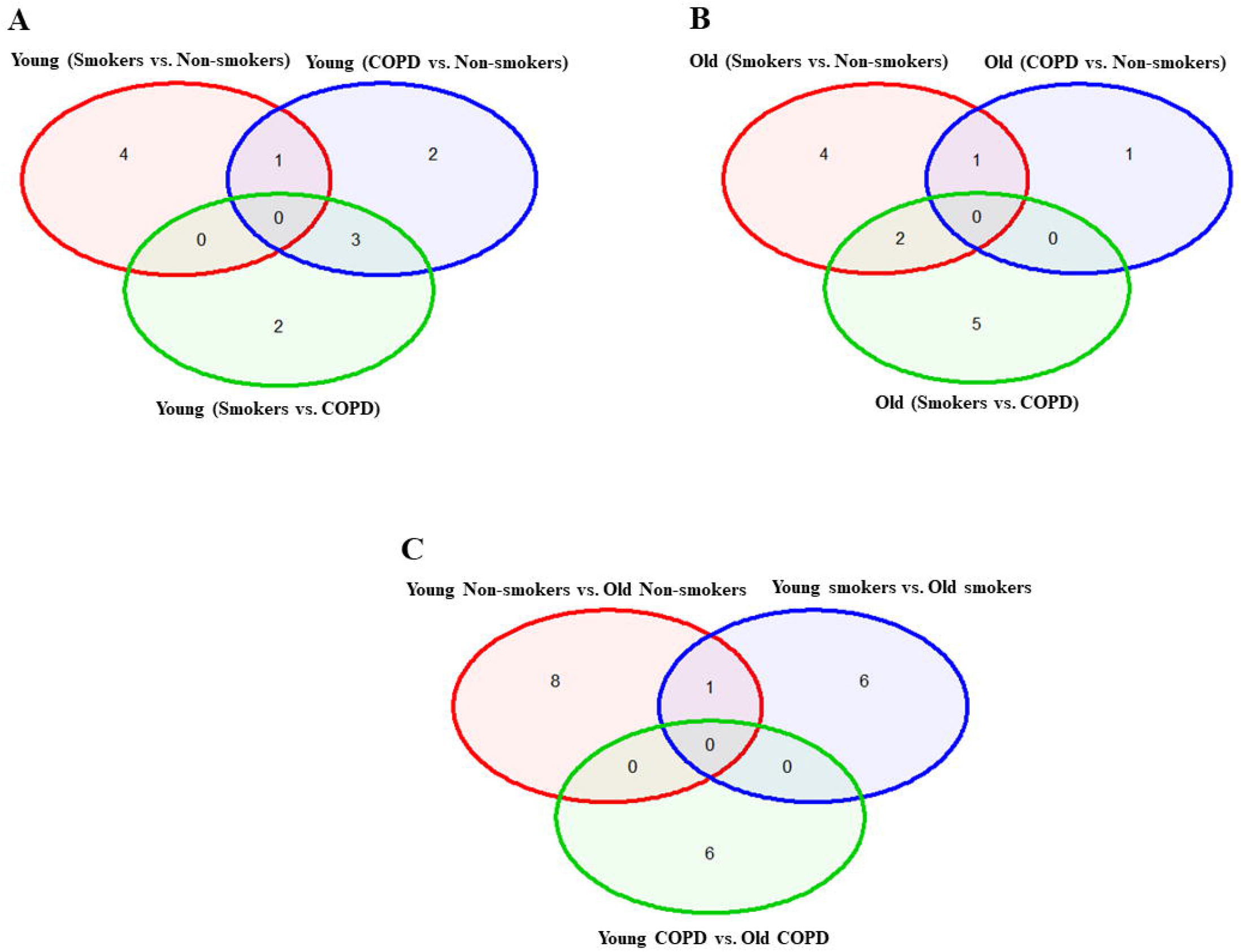
Venn diagram showing the number of altered mRNA transcripts analyzed by NanoString. Venn diagram representing the number of gene changes among Non-smokers, smokers and COPD, which were further divided for pairwise comparisons into A) young vs young, B) old vs old, and C) young vs old aged subjects. Lung RNA was isolated, processed and analyzed by NanoString. Normalized gene expressions were used for all the comparisons.

**Figure 3.**
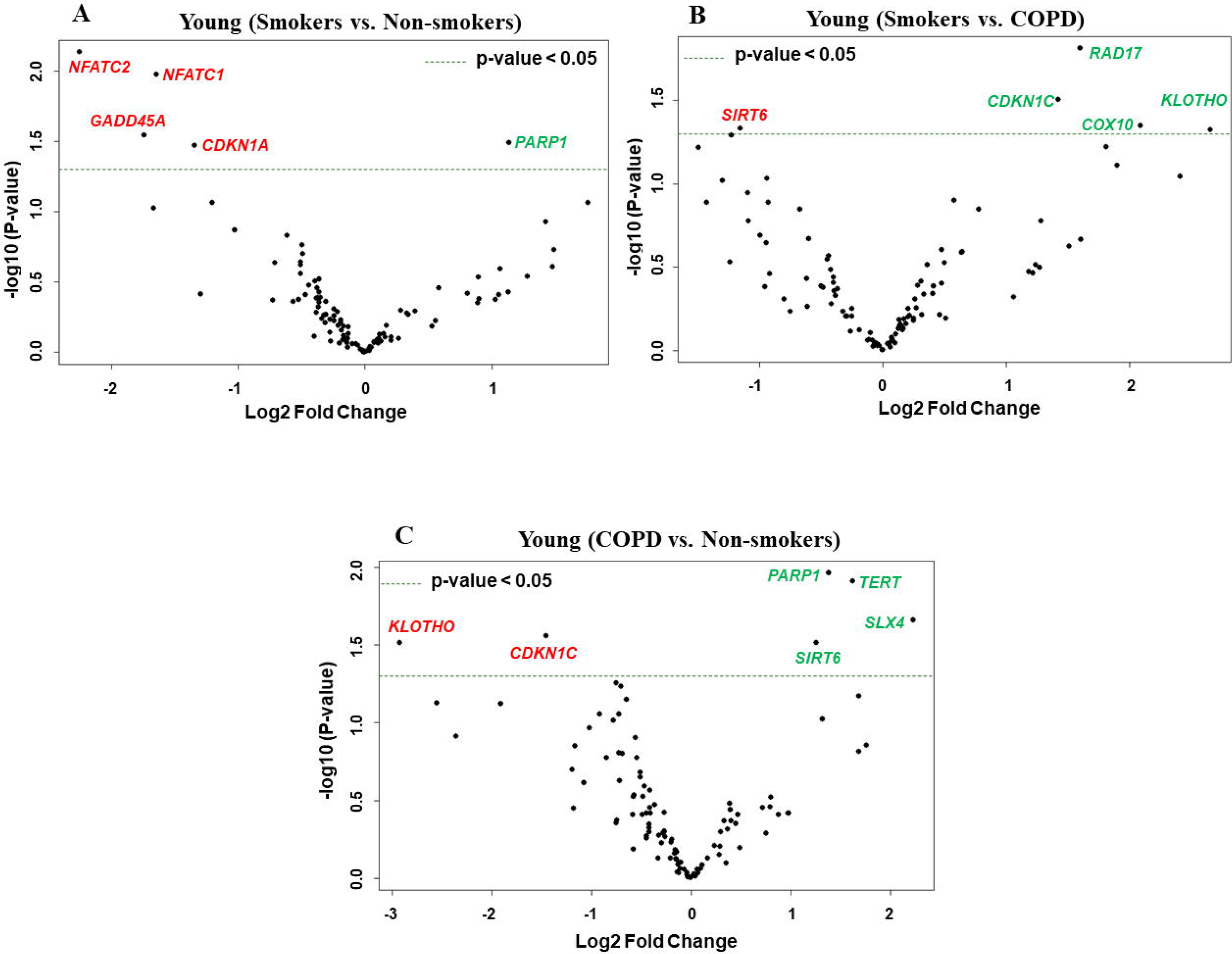
Volcano plots showing differentially expressed genes related to mitochondrial biogenesis and function, telomere replication and cellular senescence genes among non-smokers, smokers and COPD subjects. Altered genes in comparisons between A) Young (Smokers vs. Non-smokers). B) Young (Smokers vs. COPD). C) Young (COPD vs. Non-smokers). The genes differentially expressed in each comparison are indicated in green (increased) and red (decreased) colors. The green dotted horizontal line indicates the significance threshold of p-values from comparisons at *P* < 0.05. The Benjamini-Hochberg procedure was further used to adjust the p-values to control the false discovery rate at 5%.

### Differentially expressed genes in old non-smokers versus old smokers versus old COPD groups

Here we analyzed differentially expressed transcript levels among old non-smokers vs. old smokers, old smokers vs. old COPD and old non-smokers vs. old COPD groups. Out of 7 genes, we found 3 genes *IGF1, COX18* and *RIF1* were decreased and remaining 4 genes *NFATC1, NFATC2, RAD17* and *PCNA* were increased in old smokers as compared to old non-smokers group (**Figures 2B and 4A**). The transcript levels of *IGF1, PARP1, PTEN, NBN, HSPD1* and *RIF1* were decreased and *GAR1* was increased in old smokers as compared to old COPD group (**Figures 2B and 4B**). Only 2 genes were affected among old non-smokers and old COPD group; *RPA2* and *PCNA* were increased in old COPD as compared to old non-smokers group (**Figures 2B and 4C**). Overall, a total of 3 genes as mentioned above (*IGF1, RIF1* and *PCNA*) were among the common targets that was found differentially expressed in old smokers as compared to old non-smokers and old COPD groups.

**Figure 4.**
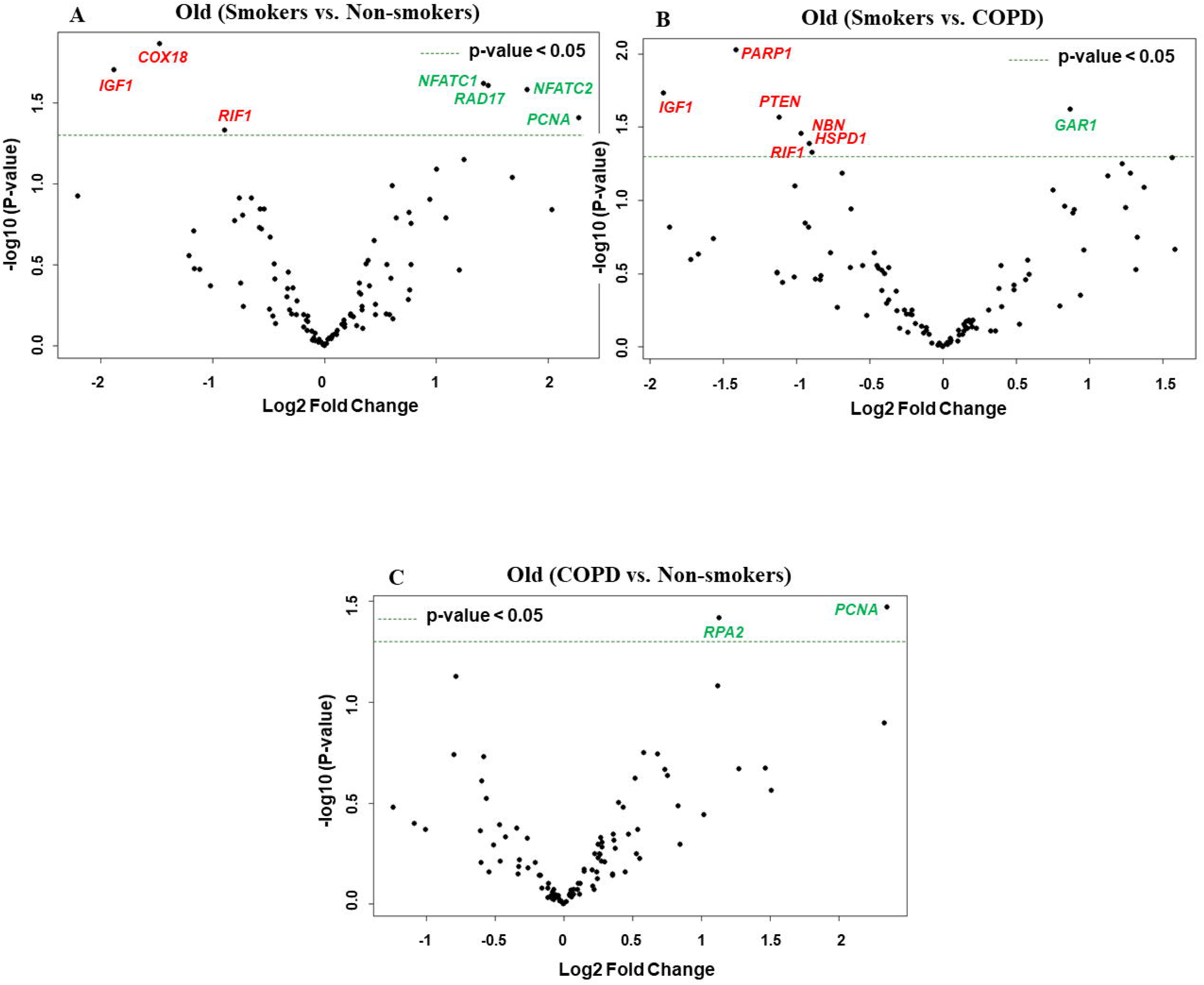
Volcano plots showing differentially expressed genes related to mitochondrial biogenesis and function, telomere replication and cellular senescence genes among non-smokers, smokers and COPD subjects. Altered genes in comparisons between A) Old (Smokers vs. Non-smokers). B) Old (Smokers vs. COPD). C) Old (COPD vs. Non-smokers). The genes differentially expressed in each comparison are indicated in green (increased) and red (decreased) colors. The green dotted horizontal line indicates the significance threshold of p-values from comparisons at *P* < 0.05. The Benjamini-Hochberg procedure was further used to adjust the p-values to control the false discovery rate at 5%.

### Altered gene expression levels in young and old non-smokers versus smokers versus COPD groups

We then analyzed differentially expressed genes among young non-smokers vs. old non-smokers, young smokers vs. old smokers and young COPD vs. old COPD pairwise comparisons. Transcript levels across different age groups were performed to better understand, whether age factor influences the measured outcomes in the current study. Accordingly, we found that 9 genes were significantly elevated in young non-smokers as compared to old non-smokers group. The following are the genes that were increased among young non-smokers (*PCNA, NFATC2, ACD, GSK3β, HAT1, UCP2, CDKN1A, CDKN1C and SIRT1*) as compared to old non-smokers group (**Figures 2C and 5A**). Although, young smokers show increased transcript levels of *PARP1, UCP3* and *E2F1* genes but decreased levels of *NFATC1, NFATC2, MYC* and *GADD45A* as compared to old smokers group as seen in **Figures 2C and 5B**. Additionally, 2 genes *TNSK2* and *PTEN* were decreased in young COPD as compared to old COPD group. The transcript levels of *GAR1, TERT, H2AX* and *FEN1* tend to increase in younger COPD as compared to old COPD group (**Figures 2C and 5C**). Interestingly, we noted that transcript levels of *NFATC2* was decreased in lungs of old non-smokers, but increased in lungs of old smokers.

**Figure 5.**
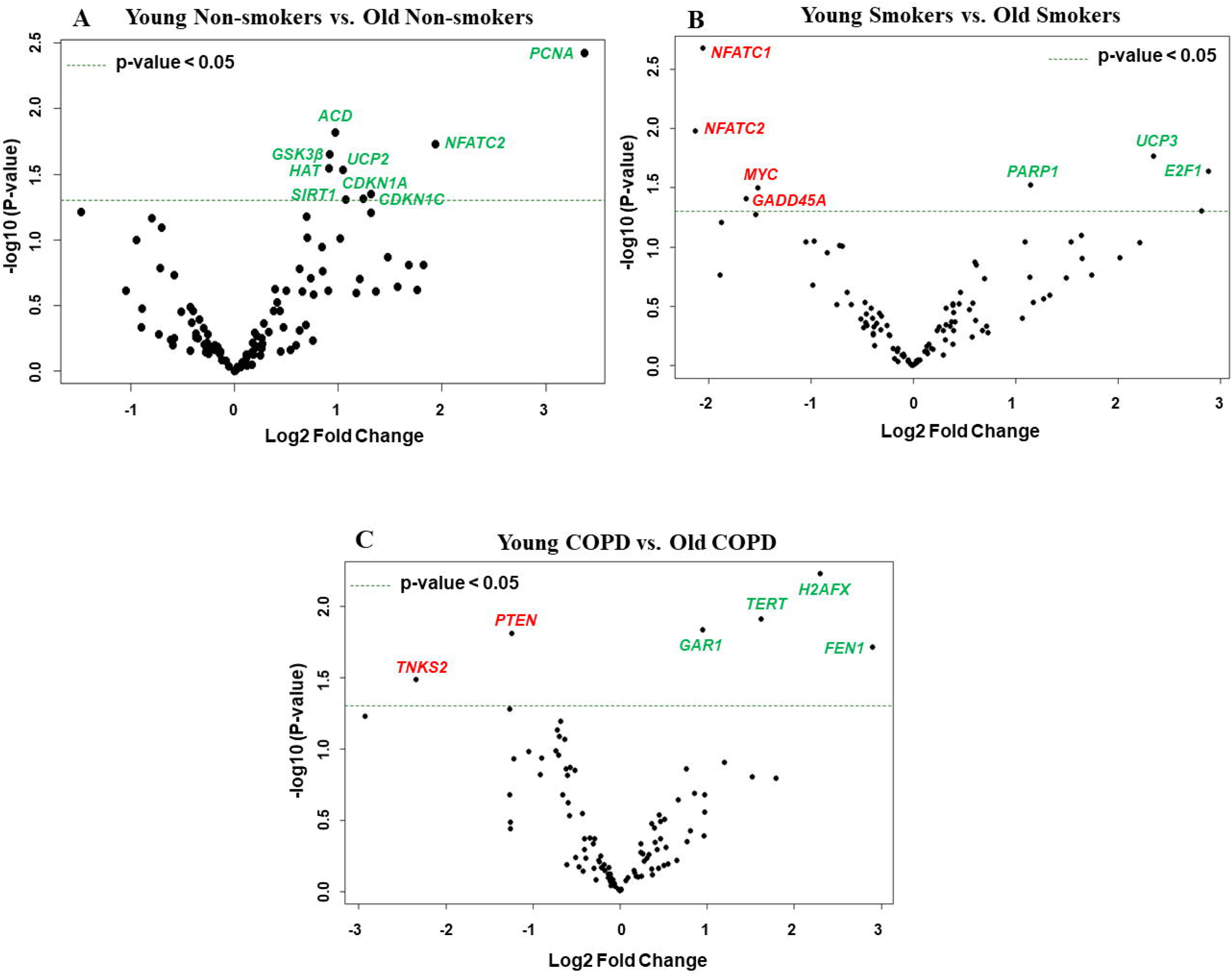
Volcano plots showing differentially expressed genes related to mitochondrial biogenesis and function, telomere replication and cellular senescence genes among non-smokers, smokers and COPD subjects. Altered genes in comparisons between A) Non-smokers (Young vs. Old). B) Smokers (Young vs. Old). C) COPD (Young vs. Old). The genes differentially expressed in each comparison are indicated in green (increased) and red (decreased) colors. The green dotted horizontal line indicates the significance threshold of p-values from comparisons at *P* < 0.05. The Benjamini-Hochberg procedure was further used to adjust the p-values to control the false discovery rate at 5%.

### Combined analysis of differentially expressed genes among non-smokers, smokers and COPD groups

We performed grouped analysis of differentially expressed transcripts (comparisons without considering the age factor (young or old) among non-smokers, smokers and patients with COPD groups. Data from young and old that belong to same experimental groups were combined (young nonsmokers with old non-smokers group, young smokers with old smokers group and young COPD and old COPD group; n=8/group, as given in **Figures 6 and 7A**). Results indicated that smokers show decreased *FOXO1* and increased *RAD17* levels as compared to non-smokers group (**Figure 8A**). While decreased *PARP1* and increased *RAD17* levels were observed in smokers as compared to COPD group (**Figure 8B**). In patients with COPD *KLOTHO* gene was decreased and *PARP1* and *SLX4* genes were increased as compared to non-smokers group (**Figure 8C**). Furthermore, these comparisons revealed that smokers have significantly higher levels of *RAD17* expression compared to both non-smokers and COPD groups. Whereas, COPD patients showed significantly higher levels of *PARP1* as compared to both non-smokers and smokers groups.

**Figure 6.**
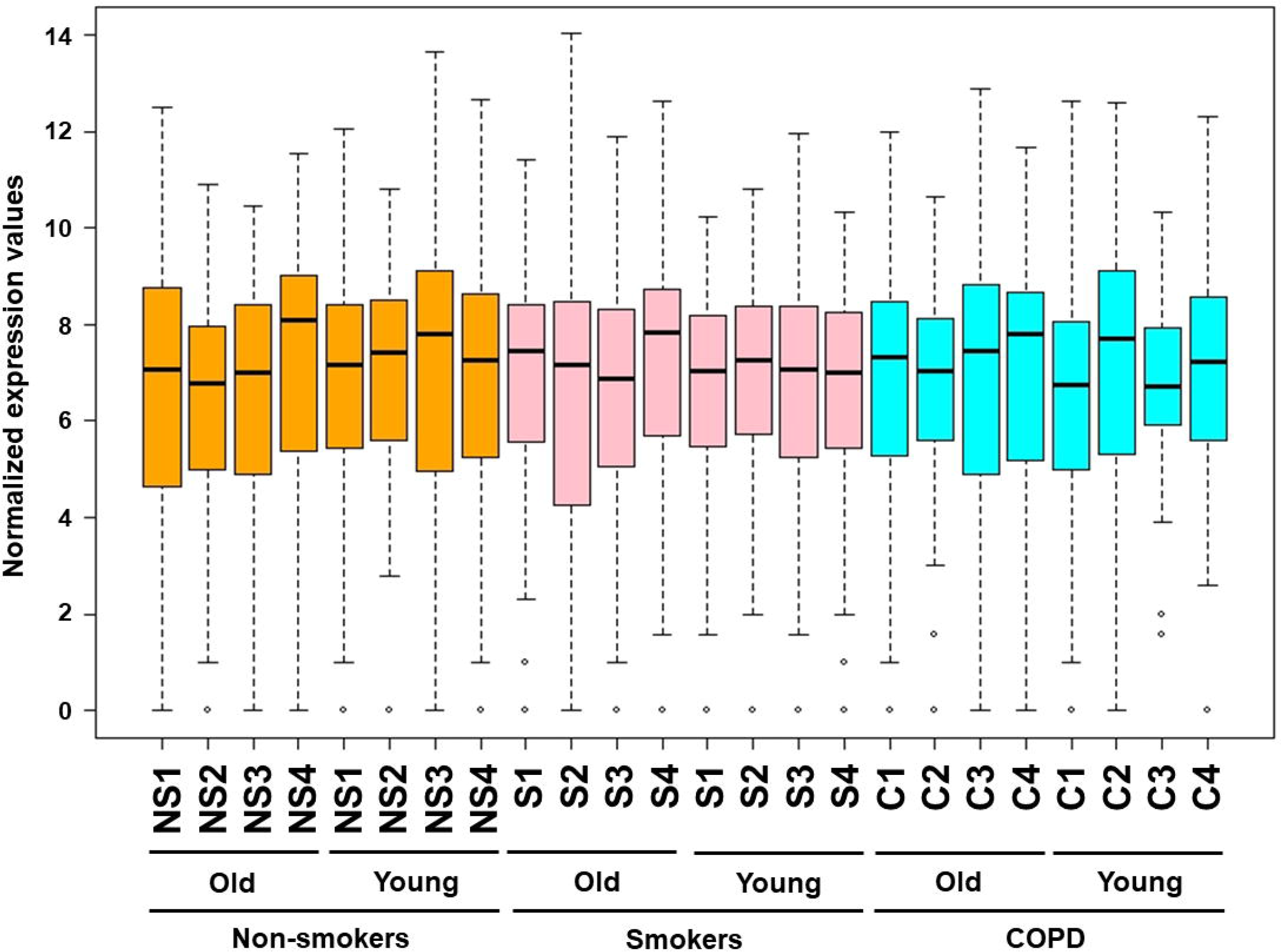
Boxplot analysis of normalized mRNA transcript analyzed by NanoString. Boxplot shows distribution of normalized gene expression levels in combined subjects from non-smokers, smokers and COPD subjects.

**Figure 7.**
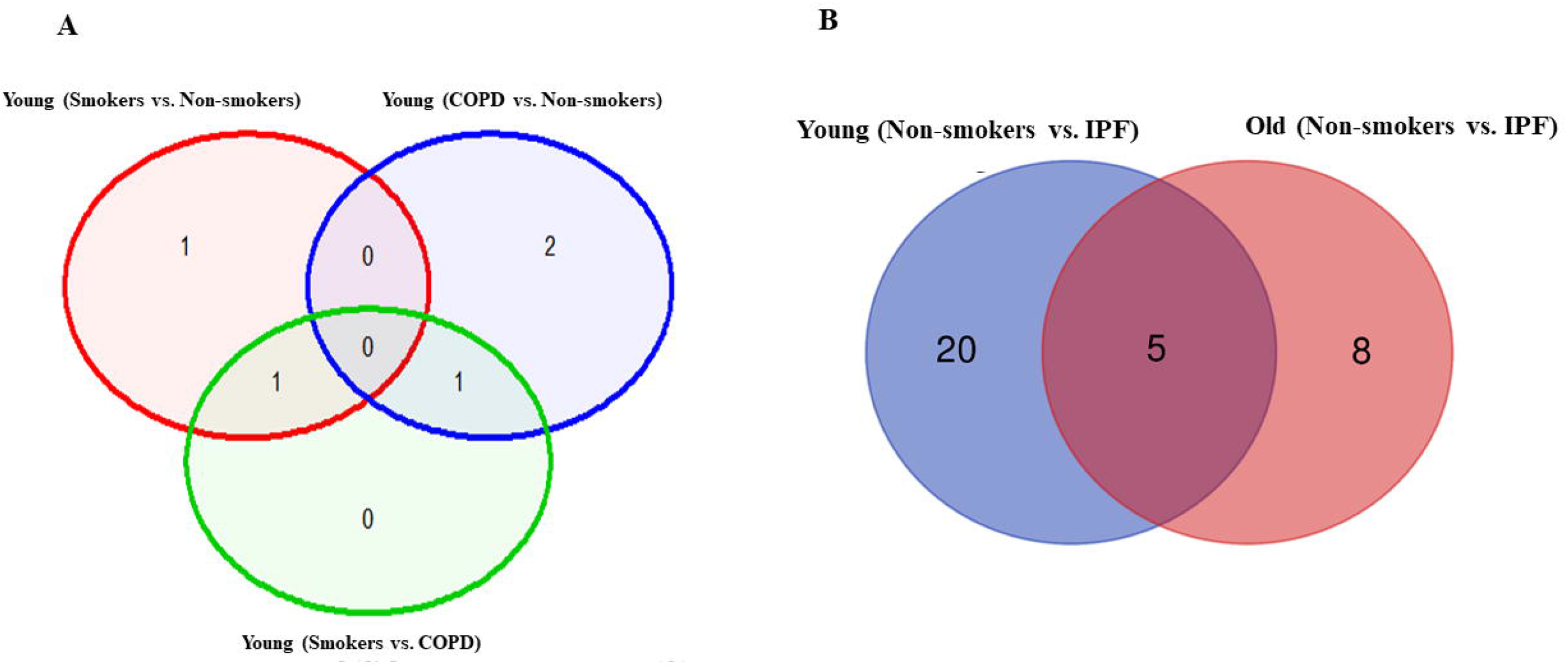
Venn diagram showing the number of altered mRNA transcripts analyzed by NanoString. Venn diagram representing the number of gene changes among A) Non-smokers, smokers and COPD. B) Non-smokers and IPF groups. Lung RNA was isolated, processed and analyzed by NanoString. Normalized gene expressions were used for all the comparisons.

**Figure 8.**
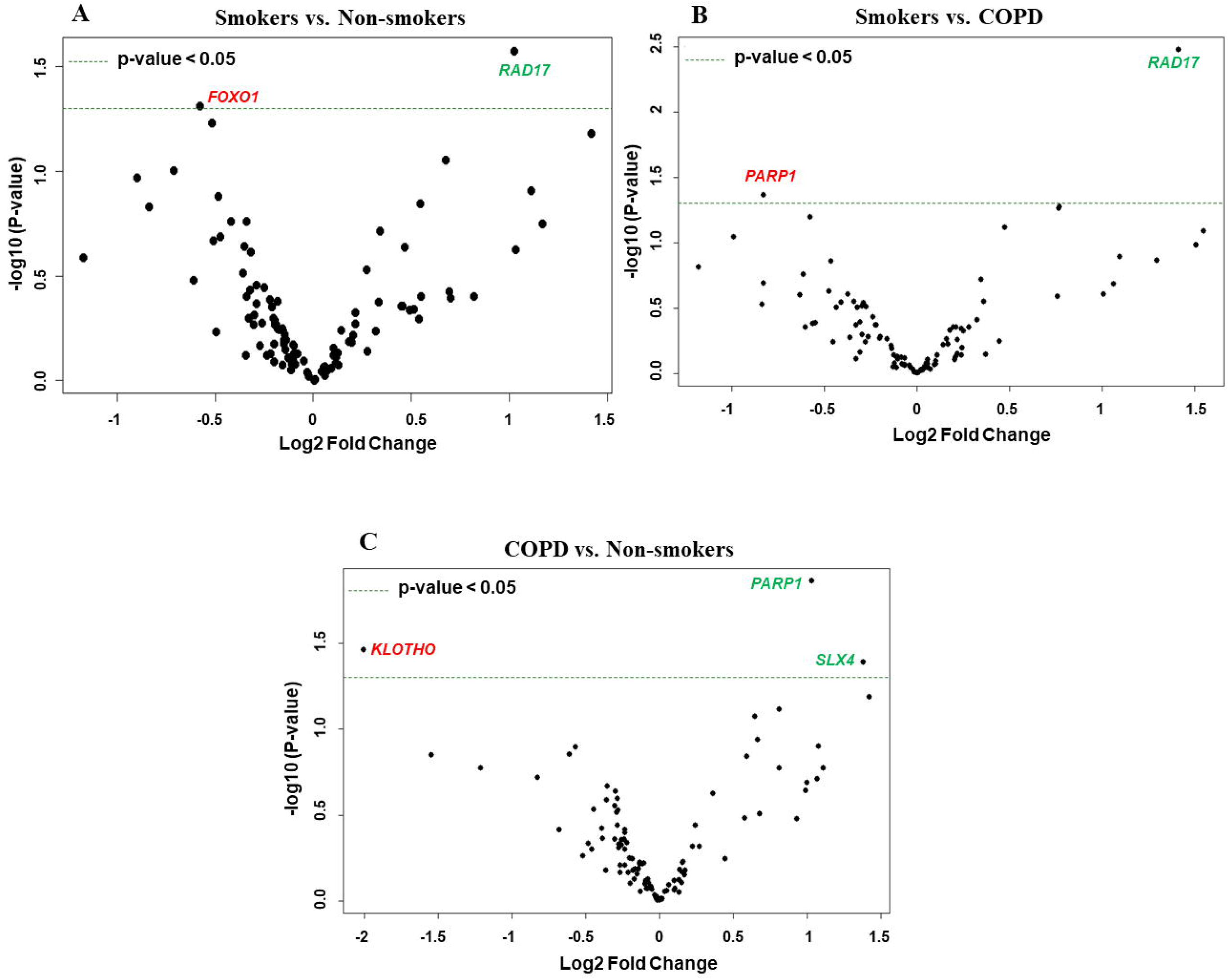
Volcano plots showing differentially expressed genes related to mitochondrial biogenesis and function, telomere replication and cellular senescence genes among non-smokers, smokers and COPD subjects. Altered genes in comparisons between A) Smokers vs. Non-smokers. B) Smokers vs. COPD. C) COPD vs. Non-smokers. The genes differentially expressed in each comparison are indicated in green (increased) and red (decreased) colors. The green dotted horizontal line indicates the significance threshold of p-values from comparisons at *P* < 0.05. The Benjamini-Hochberg procedure was further used to adjust the p-values to control the false discovery rate at 5%.

### Altered gene expression levels in young and old non-smokers versus IPF groups

Pairwise analysis of young non-smokers vs young IPF showed 25 significantly altered genes, which were given in **Table 2**. Comparisons between old non-smokers and old IPF showed 13 significantly altered genes, as listed in table 3 along with their observed level of significance. The gene comparisons among these groups were given in **Figure 7B**.

### Altered gene expression levels in young and old COPD versus IPF groups

As detailed above both COPD and IPF are chronic age related diseases that severely alter lung function and share certain common features for the disease occurrence and progression. Here, we compared the altered gene levels related to the same pathways among COPD and IPF subjects as detailed above. There was no change in any of the genes analyzed in comparisons between young IPF (n=3) and old IPF (n=13). While, 16 genes were found to be altered in the comparisons between young (COPD vs IPF), as indicated in **Table 4**. A total of 6 genes were altered in comparisons between old COPD vs old IPF) as shown in **Table 5**.

**Table 4.** Altered genes in comparisons between young COPD and young IPF subjects

| List of genes that were found to be increased in young IPF compared to young COPD |  |
| --- | --- |
| Genes | p values |
| ALDH1A3 | 0.004086 |
| COX10 | 0.019541 |
| IGF1 | 0.024304 |
| NOX4 | 0.001143 |
| RAD50 | 0.014307 |
| TNKS2 | 0.006571 |
| UCP3 | 0.043174 |
| List of genes that were found to be decreased in young IPF compared to young COPD |  |
| AIP | 0.041142 |
| ATP5O | 0.02557 |
| CDK2 | 0.042857 |
| ERCC1 | 0.039613 |
| FIS1 | 0.011772 |
| GADD45B | 0.004059 |
| ID1 | 0.009297 |
| RHOT2 | 0.022329 |
| SP1 | 0.013991 |

**Table 5.** Altered genes in comparisons between old COPD and old IPF subjects

| List of genes that were found to be decreased in old IPF compared to old COPD |  |
| --- | --- |
| Genes | p values |
| ATP5L | 0.033729 |
| CDKN1B | 0.026684 |
| CDKN1C | 0.010141 |
| GADD45B | 0.021874 |
| SIRT2 | 0.018325 |
| TSC2 | 0.033306 |

### Gene expression analysis of differentially expressed targets

Some of the differentially expressed mRNA targets predicted using Nanostring were selected for qPCR study. As given in the **Figure 9** the expression trends of these selected genes matches with the trend observed using the NanoString mRNA analysis with varied levels of fold changes. Combined gene analysis for *PARP1* was also given, and matches with the trend are given in **Figure 8**. The results clearly indicate that the genes validated using qPCR are in agreement and significant across all the pairwise comparisons made.

**Figure 9.**
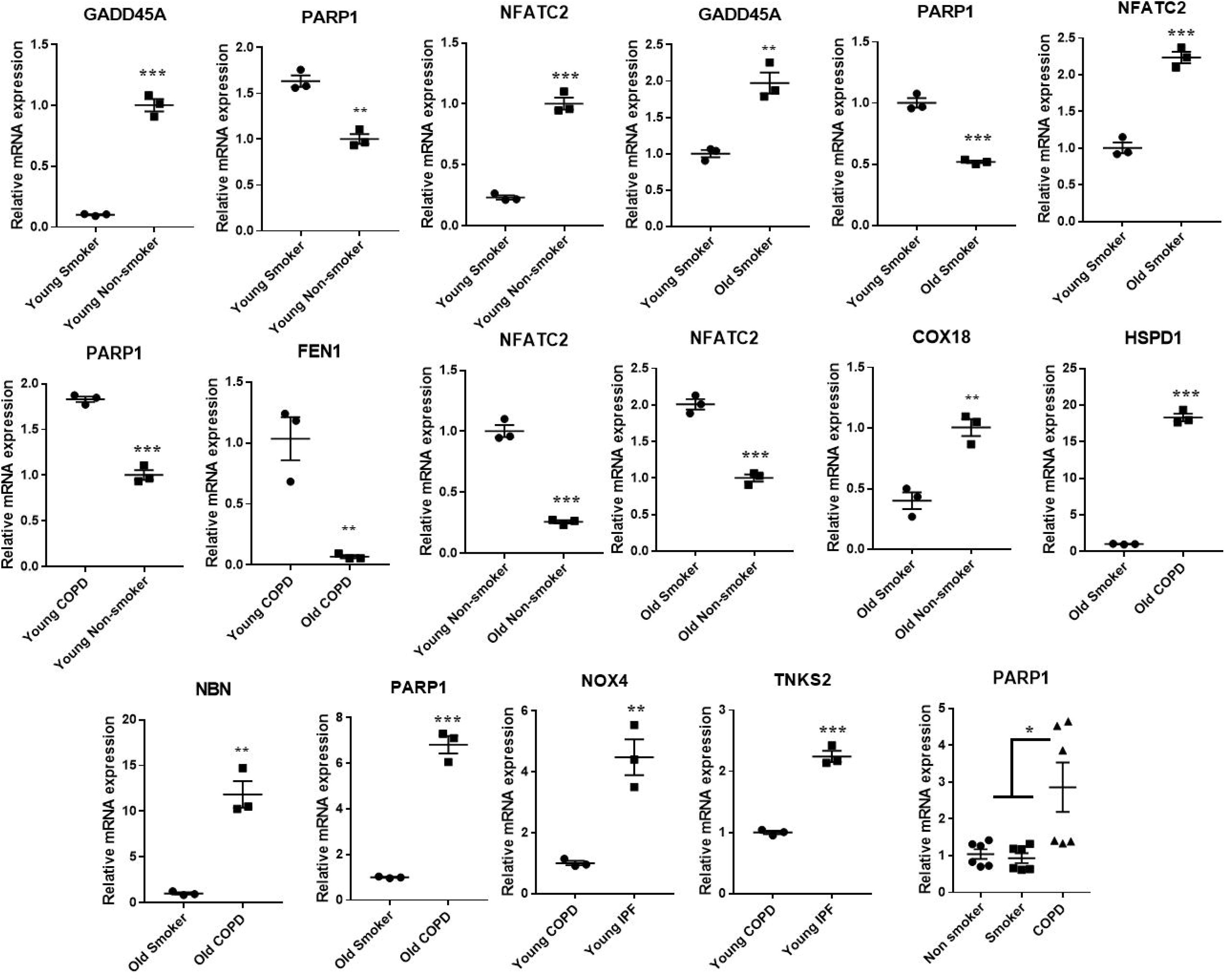
Quantitative PCR validation of the selected genes, which were found to significantly and differentially altered across various pairwise comparisons. The values were deduced based 2-ΔΔCt method. The genes represented were found to be significant in their pairwise comparisons (p0<05). Student t-test was used to compare the level of significance in pairwise comparisons, while ANOVA was used for multiple comparisons.

### Protein expression levels of crucial SARS-CoV-2 targets in the lung homogenates

Western blots analysis (**Figures 10 and 11**) revealed a significant increase in the protein levels of TMPRSS2 protease (which plays a crucial role in the processing of the SARS-CoV-2 proteins) in COPD subjects as compared to smokers and non-smokers. Similarly, the levels of another important protease furin a spike protein, was also increased in smokers and COPD subjects, with a significant expression in COPD subjects. ACE2, which is considered crucial for the SARS-CoV2 binding as a receptor, was found to increase in smokers as compared to the rest of the groups. The samples were also further probed for the expression of DPP4, another crucial protein which plays an important role in MERS-CoV binding. The DPP4 expression was found to be significantly higher in smokers as compared to COPD and non-smokers. These results indicate that the levels of proteases, which aid in the processing and binding of the viral spike proteins were highly expressed in COPD and smokers. The expression results showed varied protein intensities in smokers and COPD for TMPRSS2 and DPP4 in the lungs, which may suggest a varied effect of virus entry/susceptibility based on specific cells in smokers and COPD/IPF subjects.

**Figure 10.**
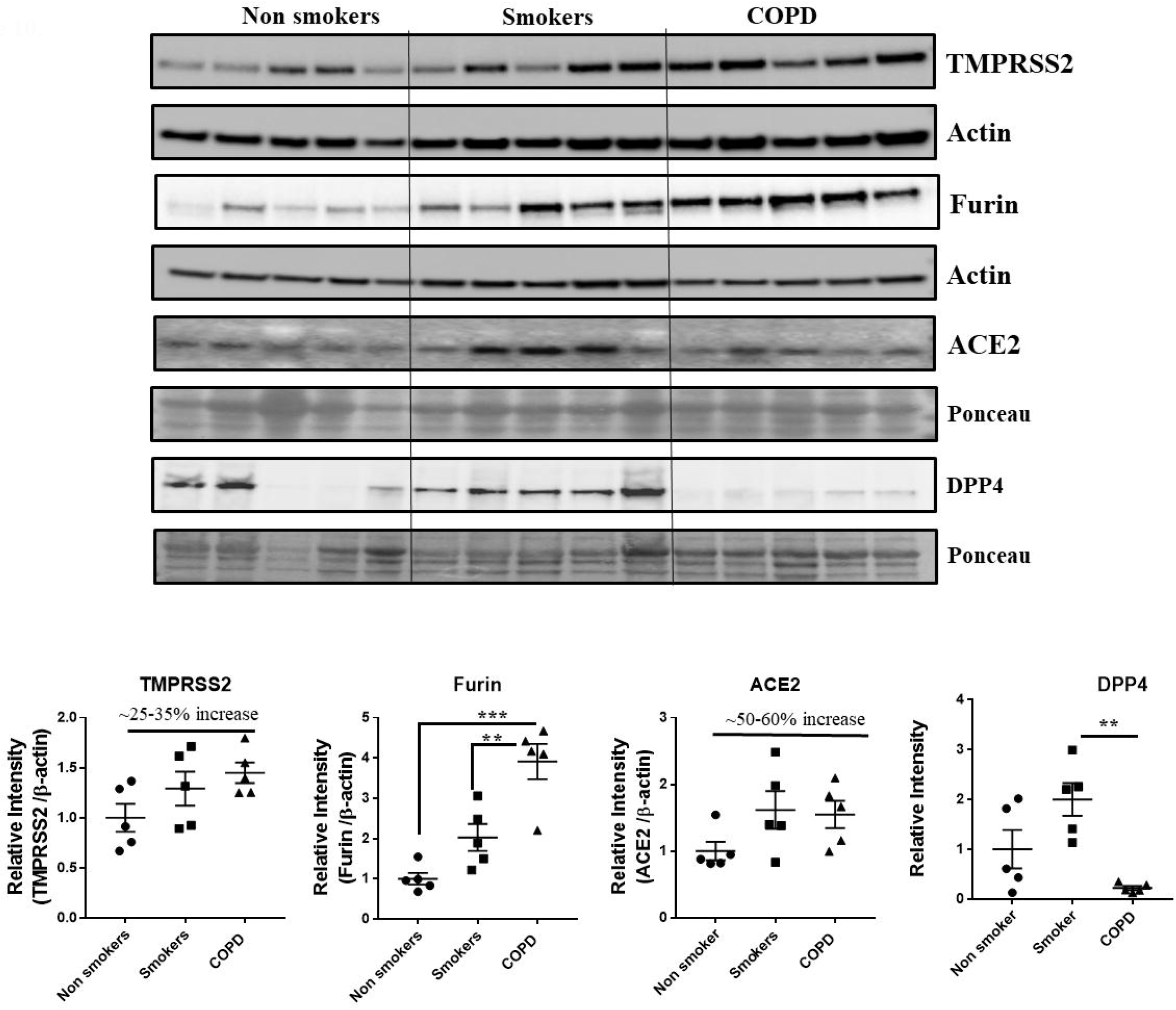
Western blot analysis of the crucial targets involved in COVID19 in non-smokers, smokers and COPD subjects. Five samples per groups were used to probe for the TMPRSS2, furin, ACE2 and DPP4. Data were shown as mean ± SEM (n = 5/group). Level of significance were indicated as * P < 0.05 and ** P < 0.01 across the groups.

**Figure 11.**
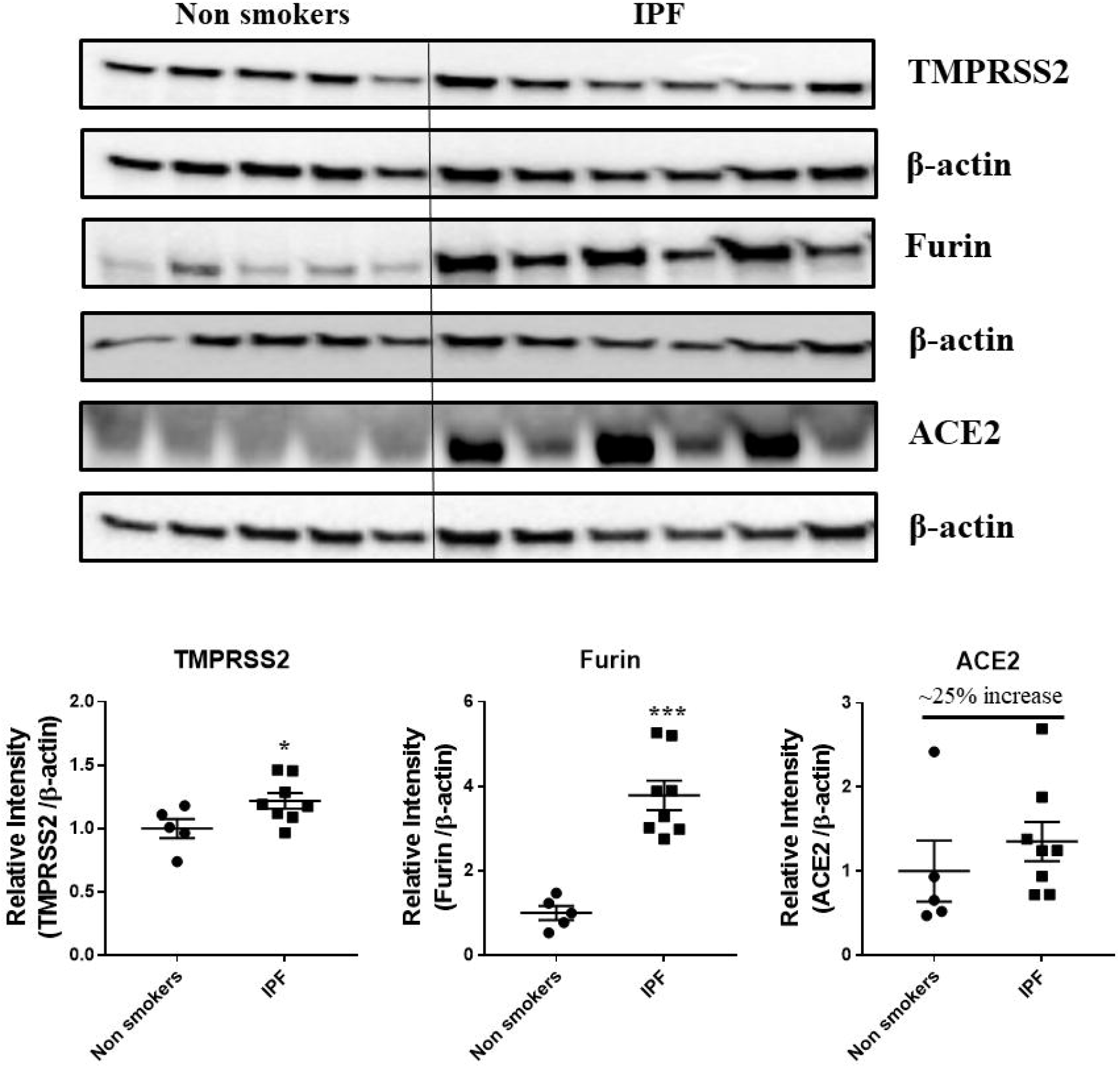
Western blot analysis of the crucial targets involved in COVID19 in non-smokers and IPF subjects. Five samples per groups were used to probe for the TMPRSS2, furin and ACE2. Data were shown as mean ± SEM (n = 5/group). Level of significance were indicated as * P < 0.05 and ** P < 0.01 across the groups.

## Discussion

Aging is a major contributor for the decline in lung function and as most of the COPD are old aged, so there is a need to define the role of aging influence in COPD. On the other hand, smoking is one of the major contributing factors for the development of COPD. COPD, is commonly observed in the older subjects compared to younger ones. Nonetheless there are growing evidences of young subjects with COPD, which needs a thorough and careful phenotypic characterization for potential markers to understand the cause and progression of disease. The current study examined the changes in the gene expression in the normal lungs of non-smokers and smokers and patients with COPD/IPF. The study included age as an influencing factor apart from the lung disease status. The extracted RNA was processed and analyzed using the sophisticated NanoString nCounter analysis platform. The NanoString has several advantages in simultaneous estimation of the several gene levels in a single sample with low amounts of sample input ^43,44^. We have successfully used this platform to report the changes in gene levels using different gene set panels in our earlier studies ^17,29^. In the current study, the custom designed panel included genes from three different pathways addressing the mitochondrial biogenesis and function, telomere function and cellular senescence, which play a major role in the lung inflammation and COPD development.

Most of the chronic diseases like COPD are associated with the mitochondrial dysfunctions. Several reports claim the causal role of mitochondrial dysfunctions in the initiation and progression of smoking associated COPD ^45-47^. We have previously reported the presence of mitochondrial dysfunctions in CS-induced lung damage models and in human lungs ^11^. Further, we along with others have reported that telomere dysfunction is also seen in the COPD patients and smoking plays as crucial role in influencing the telomere genes ^10,14,21^. Assessment of all these three pathway related genes in the same subjects throws light on the involvement and coordination of complex processes in smoking-related chronic lung disease such as COPD.

Pairwise comparisons revealed that a total of 21 genes were altered among the young and old subjects. Among them some of the genes were earlier reported to be altered in smoker and COPD patients. *CDKN1A* (p21) which is a cell dependent kinase (CDK), plays a vital role in the cellular senescence and proliferation was reported to be increased in smokers and COPD subjects ^48^. Further, p21 disruption attenuated CS-induced lung inflammation in mice ^49^. In the current study, there was a reduced expression in the p21 levels in the young smokers compared to NS in accordance with the previous reports, where *CDKN1C* (p57), along with p21 was decreased in aging lungs of mice ^50^.

Among the other important targets *KLOTHO, SIRT6, PARP1* and *PCNA* were altered in the current study. COPD patients has reduced KLOTHO expression as compared to non-smokers, in accordance with previous reports ^51^. In similar lines *PARP1* was also elevated in the COPD subjects compared to smokers and non-smokers, as reported earlier ^52,53^. We have reported that *TERT* levels were altered in mice (young and old) exposed to chronic CS ^20^. Accordingly, the current results demonstrates that *TERT* levels were significantly increased in COPD patients, but this gene may be influenced in a different way in humans, as younger COPD patients has higher *TERT* levels compared to older ones. Nuclear factor of activated T cells c2 (*NFATC2*) levels were significantly higher in aged smokers as compared to younger ones. Earlier reports claim that *NFATC2* levels were increased by nicotine/smokers ^54^. It was also reported that *NFATC2* enhances tumor-initiating phenotypes in lung adenocarcinoma ^55^. These observations were opposite in non-smokers, as they age the levels of *NFATC2* were decreased. This suggests that *NFATC2* may be used as a potential marker related to CS-induced lung damage. Among the other important genes that were altered and are crucial in the maintenance of these three pathways are ACD, which is related to *TPP1* gene and coordinates with its function was elevated in COPD ^21,56^. *FEN1* could be a novel biomarker for COPD which was increased in young COPD as compared to old. Prior studies showed mutation in *FEN1* linking lung cancer progression in an age-dependent manner in mice exposed to benzo[α]pyrene which is present in tobacco smoke ^21,57^.

Pairwise comparisons between non-smokers and IPF subjects revealed changes in some of the important genes like *PARP1, PCNA, FEN1, CDKN1B, NFATC2* and *GADD45B*, as discussed in above comparisons. However, the directionality of the changes varied between groups, which may be attributed to the small sample size in the study and the heterogeneity in the samples used. In view of the growing interests, comparisons were also made between the expression profiles of the young IPF and old IPF with their age matched COPD subjects. Interestingly some of the well characterized genes in IPF like *NOX4, TNKS2* was increased in the young IPF as compared to the young COPD patients ^58,59^. Mitophagy is a well-known phenomenon occurring in both COPD and IPF and regulates the mitochondrial related damage response towards the disease ^45,60^. Genes participating in the mitochondrial dynamics and other quality control mechanisms like *FIS1* and *RHOT2* were found to be decreased in young IPF compared to their age matched COPD subjects. *ERCC1* (Excision Repair Cross-Complementation Group 1) was also found to be high in young COPD as compared to IPF. Earlier reports claim that *ERCC1* gene is strongly associated with COPD subjects ^61^. Some of the common gene targets like *GADD45B* needs further attention and characterization especially in the chronic lung diseases like COPD and IPF. Recent biomarker identification study indicated *GADD45B* in their list of genes that can be targeted in chronic diseases like asthma, IPF and COPD ^6^.

Several studies indicate that the patients with underlying chronic disease conditions like diabetes, hypertension and COPD are more prone to COVID19 infections and have higher chances of the hospitalization rates and mortality ^33,35,36^. Several crucial mechanisms and targets were reported to increases these susceptibility in these patients ^37^. In the current study, we have determined the expression of four such important protein targets reported to play an important role in SARS-CoV-2 COVID-19. As anticipated, COPD and IPF patients in our study have higher levels of TMPRSS2 proteins in the lungs, suggesting the ideal condition for the processing of the viral protein and attachment to its receptor ACE2. ACE2 receptor abundance expression was slightly increased, but was tend to be on higher side in smokers, COPD and IPF subjects ^62^. Furin, another crucial protease in the COVID-19 infection was also found to be higher in smokers, COPD and IPF as compared to the non-smokers. We have also assessed the levels of DPP4 in the same samples and found that DPP4 was significantly higher in the smokers as compared to non-smokers and COPD. DPP4 was found to play an important role in the entry of MERS-CoV and acts as its receptor (belonging to the similar class of beta coronaviruses) in humans, was also suggested to play an important role in COVID-19 infections ^63^. In agreement with recent reports ^38^, suggesting that smokers and COPD have higher DPP4, our findings suggest increase in DPP4 in smokers, but to our surprise we didn’t find any increase in COPD subjects. This may suggest a different mechanism in smokers and COPD, which required further studies. Nevertheless, the varied abundance of different SARS-CoV-2 COVID-19 proteins suggest cell-specific effects for viral entry in smokers and COPD/IPF subjects.

In conclusion, our study provides the novel directions in conducting the studies involving the crucial and interdependent mitochondrial, telomere and cellular senescence pathways in the same subjects in association with SARS-CoV-2 COVID-19 proteins. Whilst the study provides several differential gene expression patterns in non-smokers, smokers and COPD/IPF, there is a limitation regarding the small sample size and smoking history. Nonetheless the study is valuable in terms of approach and identifying a good number targets, which needs thorough characterizations in future studies.

## Data Availability

All data are available within the manuscript.

## Author contributions

KPM, IKS and IR designed and interpreted the experimental results. KPM performed all the experiments, DL analyzed the data and edited the manuscript, and prepared Figures 1-8. KPM prepared and edited Figures 1-11. KPM, IKS and IR edited, improvised and approved the final version of the manuscript.

## Additional Information

### Supplementary information

accompanies this paper (Supplementary Table S1, Supplementary Table S2, and Supplementary Figures S1-11).

### Data availability statement

We declare that we have provided all the data in the manuscript.

## Supplementary Tables

**Supplementary Table S1:** Pre-selected genes belonging to the mitochondrial biogenesis and function pathway with their annotations

**Supplementary Table S2:** Determined genes with raw and normalized counts based on Nano String analysis.

**Supplementary Figures S1-9**. Dot plot representation of the significantly altered genes across various groups indicated. The data were represented as the normalized counts among all the observed groups. Supplementary Figure 10-11. Full unedited gels/blots for Figures 10 and 11 (original and unprocessed) are shown.

## References

1 Skloot, G. S. The Effects of Aging on Lung Structure and Function. Clin Geriatr Med 33, 447–457, doi:10.1016/j.cger.2017.06.001 (2017).

2 Wahba, W. M. Influence of aging on lung function--clinical significance of changes from age twenty. Anesth Analg 62, 764–776 (1983).

3 Rojas, M. et al. Aging and Lung Disease. Clinical Impact and Cellular and Molecular Pathways. Ann Am Thorac Soc 12, S222–227, doi:10.1513/AnnalsATS.201508-484PL (2015).

4 Wheaton, A. G. et al. Chronic Obstructive Pulmonary Disease and Smoking Status - United States, 2017. MMWR Morb Mortal Wkly Rep 68, 533–538, doi:10.15585/mmwr.mm6824a1 (2019).

5 Gershon, A. S. et al. in D102. OPTIMIZING OUTCOMES IN COPD American Thoracic Society International Conference Abstracts A7106-A7106 (American Thoracic Society, 2019).

6 Maghsoudloo, M., Azimzadeh Jamalkandi, S., Najafi, A. & Masoudi-Nejad, A. Identification of biomarkers in common chronic lung diseases by co-expression networks and drug-target interactions analysis. Mol Med 26, 9, doi:10.1186/s10020-019-0135-9 (2020).

7 Rahman, I. & Adcock, I. M. Oxidative stress and redox regulation of lung inflammation in COPD. Eur Respir J 28, 219–242, doi:10.1183/09031936.06.00053805 (2006).

8 Birch, J., Barnes, P. J. & Passos, J. F. Mitochondria, telomeres and cell senescence: Implications for lung ageing and disease. Pharmacol Ther 183, 34–49, doi:10.1016/j.pharmthera.2017.10.005 (2018).

9 Nyunoya, T. et al. Cigarette smoke induces cellular senescence. Am J Respir Cell Mol Biol 35, 681–688, doi:10.1165/rcmb.2006-0169OC (2006).

10 Liu, L., Trimarchi, J. R., Smith, P. J. & Keefe, D. L. Mitochondrial dysfunction leads to telomere attrition and genomic instability. Aging Cell 1, 40–46, doi:10.1046/j.1474-9728.2002.00004.x (2002).

11 Passos, J. F. & von Zglinicki, T. Mitochondria, telomeres and cell senescence. Exp Gerontol 40, 466–472, doi:10.1016/j.exger.2005.04.006 (2005).

12 Lerner, C. A., Sundar, I. K. & Rahman, I. Mitochondrial redox system, dynamics, and dysfunction in lung inflammaging and COPD. Int J Biochem Cell Biol 81, 294–306, doi:10.1016/j.biocel.2016.07.026 (2016).

13 Ahmad, T. et al. Impaired mitophagy leads to cigarette smoke stress-induced cellular senescence: implications for chronic obstructive pulmonary disease. FASEB J 29, 2912–2929, doi:10.1096/fj.14-268276 (2015).

14 Morla, M. et al. Telomere shortening in smokers with and without COPD. Eur Respir J 27, 525–528, doi:10.1183/09031936.06.00087005 (2006).

15 de-Torres, J. P. et al. Telomere length, COPD and emphysema as risk factors for lung cancer. Eur Respir J 49, doi:10.1183/13993003.01521-2016 (2017).

16 Zank, D. C., Bueno, M., Mora, A. L. & Rojas, M. Idiopathic Pulmonary Fibrosis: Aging, Mitochondrial Dysfunction, and Cellular Bioenergetics. Front Med (Lausanne) 5, 10, doi:10.3389/fmed.2018.00010 (2018).

17 Povedano, J. M., Martinez, P., Flores, J. M., Mulero, F. & Blasco, M. A. Mice with Pulmonary Fibrosis Driven by Telomere Dysfunction. Cell Rep 12, 286–299, doi:10.1016/j.celrep.2015.06.028 (2015).

18 Molina-Molina, M. & Borie, R. Clinical implications of telomere dysfunction in lung fibrosis. Curr Opin Pulm Med 24, 440–444, doi:10.1097/MCP.0000000000000506 (2018).

19 Putcha, N., Drummond, M. B., Wise, R. A. & Hansel, N. N. Comorbidities and Chronic Obstructive Pulmonary Disease: Prevalence, Influence on Outcomes, and Management. Semin Respir Crit Care Med 36, 575–591, doi:10.1055/s-0035-1556063 (2015).

20 Rashid, K., Sundar, I. K., Gerloff, J., Li, D. & Rahman, I. Lung cellular senescence is independent of aging in a mouse model of COPD/emphysema. Sci Rep 8, 9023, doi:10.1038/s41598-018-27209-3 (2018).

21 Ahmad, T. et al. Shelterin Telomere Protection Protein 1 Reduction Causes Telomere Attrition and Cellular Senescence via Sirtuin 1 Deacetylase in Chronic Obstructive Pulmonary Disease. Am J Respir Cell Mol Biol 56, 38–49, doi:10.1165/rcmb.2016-0198OC (2017).

22 Yao, H. et al. SIRT1 protects against emphysema via FOXO3-mediated reduction of premature senescence in mice. J Clin Invest 122, 2032–2045, doi:10.1172/JCI60132 (2012).

23 Yao, H. & Rahman, I. Role of histone deacetylase 2 in epigenetics and cellular senescence: implications in lung inflammaging and COPD. Am J Physiol Lung Cell Mol Physiol 303, L557–566, doi:10.1152/ajplung.00175.2012 (2012).

24 Cho, W. K., Lee, C. G. & Kim, L. K. COPD as a Disease of Immunosenescence. Yonsei Med J 60, 407–413, doi:10.3349/ymj.2019.60.5.407 (2019).

25 Lee, H. C., Lu, C. Y., Fahn, H. J. & Wei, Y. H. Aging-and smoking-associated alteration in the relative content of mitochondrial DNA in human lung. FEBS Lett 441, 292–296, doi:10.1016/s0014-5793(98)01564-6 (1998).

26 Saretzki, G., Murphy, M. P. & von Zglinicki, T. MitoQ counteracts telomere shortening and elongates lifespan of fibroblasts under mild oxidative stress. Aging Cell 2, 141–143, doi:10.1046/j.1474-9728.2003.00040.x (2003).

27 Tchkonia, T. & Kirkland, J.L. Aging, Cell Senescence, and Chronic Disease: Emerging Therapeutic Strategies. JAMA 320, 1319–1320, doi:10.1001/jama.2018.12440 (2018).

28 Fujii, S. et al. Insufficient autophagy promotes bronchial epithelial cell senescence in chronic obstructive pulmonary disease. Oncoimmunology 1, 630–641, doi:10.4161/onci.20297 (2012).

29 Kuznar-Kaminska, B. et al. Serum from patients with chronic obstructive pulmonary disease induces senescence-related phenotype in bronchial epithelial cells. Sci Rep 8, 12940, doi:10.1038/s41598-018-31037-w (2018).

30 Bateson, M. et al. Smoking does not accelerate leucocyte telomere attrition: a meta-analysis of 18 longitudinal cohorts. R Soc Open Sci 6, 190420, doi:10.1098/rsos.190420 (2019).

31 Gardner, I. D. The effect of aging on susceptibility to infection. Rev Infect Dis 2, 801–810, doi:10.1093/clinids/2.5.801 (1980).

32 Bonafe, M. et al. Inflamm-aging: Why older men are the most susceptible to SARS-CoV-2 complicated outcomes. Cytokine Growth Factor Rev, doi:10.1016/j.cytogfr.2020.04.005 (2020).

33 Koff, W. C. & Williams, M. A. Covid-19 and Immunity in Aging Populations - A New Research Agenda. N Engl J Med, doi:10.1056/NEJMp2006761 (2020).

34 Sargiacomo, C., Sotgia, F. & Lisanti, M. P. COVID-19 and chronological aging: senolytics and other anti-aging drugs for the treatment or prevention of corona virus infection? Aging (Albany NY) 12, 6511–6517, doi:10.18632/aging.103001 (2020).

35 Zhao, Q. et al. The impact of COPD and smoking history on the severity of COVID-19: A systemic review and meta-analysis. J Med Virol, doi:10.1002/jmv.25889 (2020).

36 Emami, A., Javanmardi, F., Pirbonyeh, N. & Akbari, A. Prevalence of Underlying Diseases in Hospitalized Patients with COVID-19: a Systematic Review and Meta-Analysis. Arch Acad Emerg Med 8, e35 (2020).

37 Hoffmann, M. et al. SARS-CoV-2 Cell Entry Depends on ACE2 and TMPRSS2 and Is Blocked by a Clinically Proven Protease Inhibitor. Cell 181, 271–280 e278, doi:10.1016/j.cell.2020.02.052 (2020).

38 Seys, L. J. M. et al. DPP4, the Middle East Respiratory Syndrome Coronavirus Receptor, is Upregulated in Lungs of Smokers and Chronic Obstructive Pulmonary Disease Patients. Clin Infect Dis 66, 45–53, doi:10.1093/cid/cix741 (2018).

39 Sundar, I. K. et al. DNA methylation profiling in peripheral lung tissues of smokers and patients with COPD. Clin Epigenetics 9, 38, doi:10.1186/s13148-017-0335-5 (2017).

40 Sanchez-Salcedo, P. et al. Disease progression in young patients with COPD: rethinking the Fletcher and Peto model. Eur Respir J 44, 324–331, doi:10.1183/09031936.00208613 (2014).

41 Maremanda, K. P., Sundar, I. K. & Rahman, I. Protective role of mesenchymal stem cells and mesenchymal stem cell-derived exosomes in cigarette smoke-induced mitochondrial dysfunction in mice. Toxicol Appl Pharmacol 385, 114788, doi:10.1016/j.taap.2019.114788 (2019).

42 Sundar, I. K. et al. Genetic ablation of histone deacetylase 2 leads to lung cellular senescence and lymphoid follicle formation in COPD/emphysema. FASEB J 32, 4955–4971, doi:10.1096/fj.201701518R (2018).

43 Eastel, J. M. et al. Application of NanoString technologies in companion diagnostic development. Expert Rev Mol Diagn 19, 591–598, doi:10.1080/14737159.2019.1623672 (2019).

44 Goytain, A. & Ng, T. NanoString nCounter Technology: High-Throughput RNA Validation. Methods Mol Biol 2079, 125–139, doi:10.1007/978-1-4939-9904-0_10 (2020).

45 Ryter, S. W. et al. Mitochondrial Dysfunction as a Pathogenic Mediator of Chronic Obstructive Pulmonary Disease and Idiopathic Pulmonary Fibrosis. Ann Am Thorac Soc 15, S266–S272, doi:10.1513/AnnalsATS.201808-585MG (2018).

46 Bialas, A. J., Sitarek, P., Milkowska-Dymanowska, J., Piotrowski, W. J. & Gorski, P. The Role of Mitochondria and Oxidative/Antioxidative Imbalance in Pathobiology of Chronic Obstructive Pulmonary Disease. Oxid Med Cell Longev 2016, 7808576, doi:10.1155/2016/7808576 (2016).

47 Zhang, W. Z. et al. Association of urine mitochondrial DNA with clinical measures of COPD in the SPIROMICS cohort. JCI Insight 5, doi:10.1172/jci.insight.133984 (2020).

48 Chiappara, G. et al. The role of p21 Waf1/Cip1 in large airway epithelium in smokers with and without COPD. Biochim Biophys Acta 1832, 1473–1481, doi:10.1016/j.bbadis.2013.04.022 (2013).

49 Yao, H. et al. Disruption of p21 attenuates lung inflammation induced by cigarette smoke, LPS, and fMLP in mice. Am J Respir Cell Mol Biol 39, 7–18, doi:10.1165/rcmb.2007-0342OC (2008).

50 Park, C. W. & Chung, J. H. Age-dependent changes of p57(Kip2) and p21(Cip1/Waf1) expression in skeletal muscle and lung of mice. Biochim Biophys Acta 1520, 163–168, doi:10.1016/s0167-4781(01)00266-4 (2001).

51 Gao, W. et al. Klotho expression is reduced in COPD airway epithelial cells: effects on inflammation and oxidant injury. Clin Sci (Lond) 129, 1011–1023, doi:10.1042/CS20150273 (2015).

52 Dharwal, V. & Naura, A. S. PARP-1 inhibition ameliorates elastase induced lung inflammation and emphysema in mice. Biochem Pharmacol 150, 24–34, doi:10.1016/j.bcp.2018.01.027 (2018).

53 Hwang, J. W. et al. Cigarette smoke-induced autophagy is regulated by SIRT1-PARP-1-dependent mechanism: implication in pathogenesis of COPD. Arch Biochem Biophys 500, 203–209, doi:10.1016/j.abb.2010.05.013 (2010).

54 Frazer-Abel, A. A. et al. Nicotine activates nuclear factor of activated T cells c2 (NFATc2) and prevents cell cycle entry in T cells. J Pharmacol Exp Ther 311, 758–769, doi:10.1124/jpet.104.070060 (2004).

55 Xiao, Z. J. et al. NFATc2 enhances tumor-initiating phenotypes through the NFATc2/SOX2/ALDH axis in lung adenocarcinoma. Elife 6, doi:10.7554/eLife.26733 (2017).

56 Else, T. et al. Tpp1/Acd maintains genomic stability through a complex role in telomere protection. Chromosome Res 15, 1001–1013, doi:10.1007/s10577-007-1175-5 (2007).

57 Wu, Z. et al. High risk of benzo[alpha]pyrene-induced lung cancer in E160D FEN1 mutant mice. Mutat Res 731, 85–91, doi:10.1016/j.mrfmmm.2011.11.009 (2012).

58 Hecker, L. et al. Reversal of persistent fibrosis in aging by targeting Nox4-Nrf2 redox imbalance. Sci Transl Med 6, 231ra247, doi:10.1126/scitranslmed.3008182 (2014).

59 Liscio, P. et al. Scaffold hopping approach on the route to selective tankyrase inhibitors. Eur J Med Chem 87, 611–623, doi:10.1016/j.ejmech.2014.10.007 (2014).

60 Hara, H., Kuwano, K. & Araya, J. Mitochondrial Quality Control in COPD and IPF. Cells 7, doi:10.3390/cells7080086 (2018).

61 de Andrade, M. et al. Genetic variants associated with the risk of chronic obstructive pulmonary disease with and without lung cancer. Cancer Prev Res (Phila) 5, 365–373, doi:10.1158/1940-6207.CAPR-11-0243 (2012).

62 Leung, J. M. et al. ACE-2 expression in the small airway epithelia of smokers and COPD patients: implications for COVID-19. Eur Respir J 55, doi:10.1183/13993003.00688-2020 (2020).

63 Bassendine, M. F., Bridge, S. H., McCaughan, G. W. & Gorrell, M. D. Covid-19 and co-morbidities: a role for Dipeptidyl Peptidase 4 (DPP4) in disease severity? J Diabetes, doi:10.1111/1753-0407.13052 (2020).

